# The LeVe CPAP System for oxygen-efficient CPAP respiratory support: Development and pilot evaluation

**DOI:** 10.1101/2021.05.24.21256987

**Authors:** P. Culmer, W. Davis Birch, I. Waters, A. Keeling, C. Osnes, D. Jones, G. de Boer, R. Hetherington, S. Ashton, M. Latham, T. Beacon, T. Royston, R. Miller, A. Littlejohns, J. Parmar, T. Lawton, S. Murdoch, D. Brettle, R. Musasizi, G. Nampiina, E. Namulema, N. Kapur

## Abstract

**Background:** The COVID-19 pandemic, caused by severe acute respiratory syndrome coronavirus 2 (SARS-CoV-2), has placed a significant demand on healthcare providers (HCPs) to provide respiratory support for patients with moderate to severe symptoms. Continuous Positive Airway Pressure (CPAP) non-invasive ventilation can help patients with moderate symptoms to avoid the need for invasive ventilation in intensive care. However, existing CPAP systems can be complex (and thus expensive) or require high levels of oxygen, limiting their use in resource-stretched environments.

**Technical Development + Testing:** The LeVe (“Light”) CPAP system was developed using principles of frugal innovation to produce a solution of low complexity and high resource efficiency. The LeVe system exploits the air flow dynamics of electric fan blowers which are inherently suited to delivery of positive pressure at appropriate flow rates for CPAP. Laboratory evaluation demonstrated that performance of the LeVe system was equivalent to other commercially available systems used to deliver CPAP.

**Pilot Evaluation:** The LeVe CPAP system was tested to evaluate safety and acceptability in a group of ten healthy volunteers at Mengo Hospital in Kampala, Uganda. The study demonstrated that the system can be used safely without inducing hypoxia or hypercapnia and that its use was well tolerated by users, with no adverse events reported.

**Conclusions:** CPAP ventilation systems provide an important treatment option for COVID-19 patients. To deliver this for the high patient numbers associated with the COVID-19 pandemic, healthcare providers require resource efficient solutions. We have shown that this can be achieved through frugal engineering of a CPAP ventilation system, in a system which is safe for use and well tolerated in healthy volunteers. This approach may also benefit other respiratory conditions which often go unaddressed in LMICs for want of context-appropriate technology.

## 1. Introduction

The COVID-19 pandemic, caused by severe acute respiratory syndrome coronavirus 2 (SARS-CoV-2), has placed a significant demand on healthcare providers (HCPs) to provide respiratory support for patients with moderate to severe symptoms [1]. Emerging clinical reports indicate that Continuous Positive Airway Pressure (CPAP) non-invasive ventilation can help patients with moderate symptoms to avoid the need for invasive ventilation in intensive care [2], [3], a change to the first impression that early intubation was indicated. Regulatory authorities such as the UK MHRA and US FDA have produced guidance to support rapid development, manufacture and approval of new ventilation systems which can be produced at scale [4], [5]. However, the demand for ventilator equipment is outstripping supply through complex international supply chains. Similarly, the high patient numbers presenting in a clinical setting has placed increased burden on hospital resources and the provision of medical oxygen crucial for ventilation has faced restrictions to avoid overloading hospital systems [6].

The need to minimise oxygen consumption per patient and reduce the complexity of equipment are paramount to consider together, this has implications on adoption within different healthcare contexts. Figure 1 uses these traits to classify the types of system available for delivering CPAP in a healthcare setting. The innovation to address provision as a result of the COVID-19 pandemic has been impressive, particularly in relation to systems using pressurised oxygen (the right hand quadrants of Figure 1) typical of many healthcare systems in high income countries (HICs). For example, the UCL-Ventura device (mid-right quadrant) based on a Respironics Whisperflow [7], has been licenced in excess of 1000 times [5]. Whilst the focus of development was on rapid delivery, the final device showed improved oxygen efficiency over its initial design as a result of engineering changes. Venturi valves (bottom right quadrant) are mechanically simple, have been used extensively within healthcare settings to deliver CPAP, and are readily scalable from a manufacturing standpoint. A high pressure source of oxygen flows through the valve and entrains air, creating a flow of enriched air at a modest pressure. Different valve designs give different FiO_2_ which allows clinicians to specify an appropriate valve from a measurement of the patient’s oxygen saturation levels, with pressure in the circuit controlled using a Positive End-Expiratory Pressure (PEEP) valve design [8], [9], [10]. Shortages of these Venturi systems saw groups 3D printing such devices on humanitarian grounds to support patient care [11], [12]. However, systems in both the right-hand quadrants require a high -flow and -pressure oxygen supply, in part because this provides the energy to generate the pressure and flows within the breathing circuit, rendering them incompatible with oxygen concentrators.

**Figure 1.**
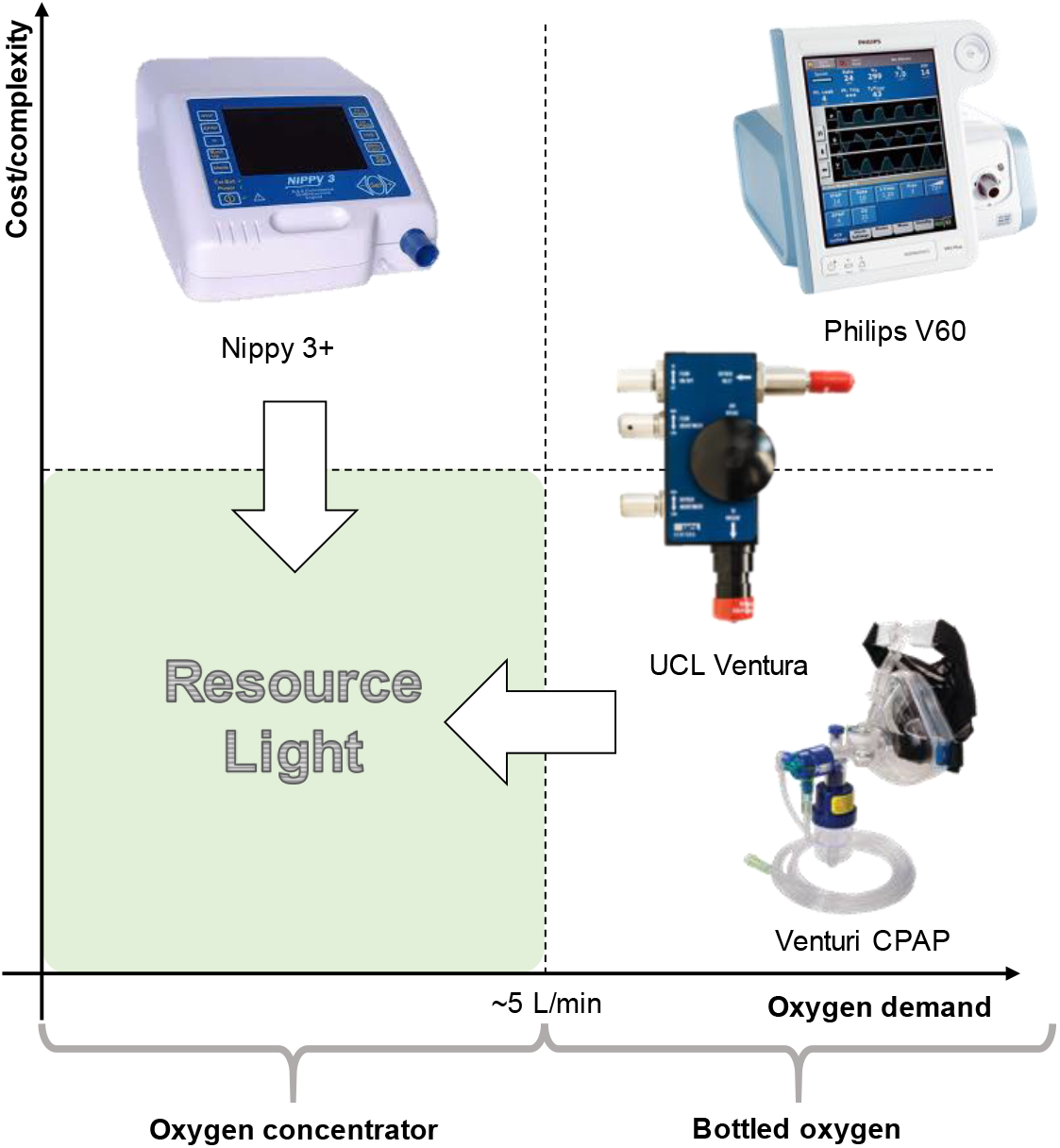
Comparison of non-invasive ventilation options for delivering CPAP

In delivering CPAP, the nature of the oxygen supply in terms of flow-per-patient and the delivery pressure is particularly relevant within low-to-middle income countries (LMICs), where limitations in healthcare infrastructure often precludes oxygen delivery from a centralised source. Instead, portable oxygen concentrators provide a sustainable, reliable and cost-effective solution in contrast to supply from compressed oxygen cylinders which require a reliable supply infrastructure and continual monitoring [13], [14]. NGOs such as UNICEF have been provisioning LMICs with oxygen concentrators for many years, recognising the need for oxygen therapies in general, which is also reflected in their inclusion in the WHO list of essential medicines [15]. The oxygen concentrators appropriate for use in LMICs typically output at low pressures and offer flow rates of 5 or 10 L/min with an oxygen concentration of ca. 95% [13]. Consequently, to deliver CPAP using oxygen concentrators, systems must fall into the left-hand side of Figure 1. Within the top left quadrant, non-invasive fan based ventilator systems including sleep apnoea systems have been used to successfully treat patients during the COVID-19 pandemic using oxygen entrained near the patient’s mask at low flow rates (ca. 5 l/min to obtain 40% FiO_2_) [16], [17].

However, existing capabilities do not currently address the bottom-left hand corner of Figure 1 – we term solutions here as *LeVe* (‘light’) – CPAP systems with low complexity and high resource efficiency – both from a design and oxygen perspective. “Frugal innovation” provides a development approach to target this region. Weyrauch et al define frugal innovation as one where products have: (i) substantial cost reduction; (ii) concentration on core functionalities; and (iii) optimised performance level [18]. Our work targets the development of systems which address the ‘*resource light’* region, a neglected but important space to consider for Global Health provision. Accordingly, here we report on the development and initial evaluation of the LeVe CPAP Blower, developed using frugal techniques to reduce the complexity of fan based systems and focus *specifically* on providing CPAP functionality.

## 2. System Development

To adapt CPAP technology into a low resource form, such that it is appropriate for use in LMICs as well as other resource-stretched situations, we hypothesise a design through examination of the working principles within existing fan-based non-invasive ventilators.

### Existing Technology

Fan based ventilators use a modulated fan to control the output pressure on a breathing circuit. The desired positive pressure is maintained through a control loop, with an internal pressure sensor used to set the appropriate fan speed. The breathing circuit is shown in Figure 2. Of critical consideration is the need to entrain oxygen close to the patient (between the mask and expiration port) to ensure high oxygen efficiency, while using an expiration port to prevent build-up of CO_2_. An example of a commercial system is the Nippy 3+ (Breas Medical Ltd), which was adopted successfully within the Leeds Teaching Hospitals NHS Trust to support COVID-19 patients. Such systems have also been demonstrated in an open-source framework with a low raw component cost although the expertise to assemble and guarantee the quality of such a system is still relatively high [18]. The functionality of these devices is typically in excess of that required to deliver CPAP therapy alone, systems typically offer more complex breathing support including bi-level positive airway pressure and automatic positive airway pressure. Since adoption into a health care setting is not only about equipment but also training of staff, this additional functionality may be disabled to ensure healthcare compliance.

**Figure 2.**
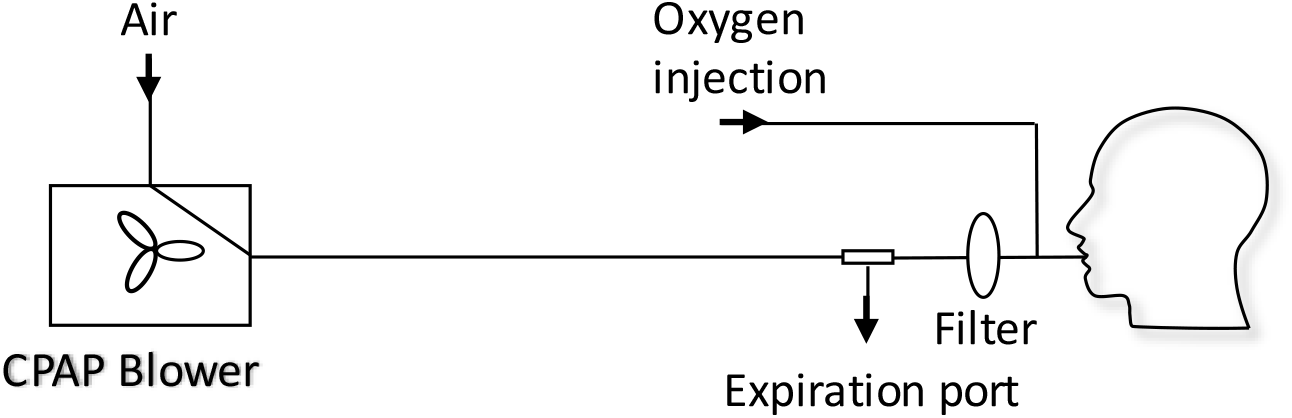
The CPAP machine is connected to an expiration port, a HEPA filter, the oxygen inlet port, and the patient mask. The expiration port is a plain hole: filtration of exhaled air before exiting into the ward could prevent aerosolization of disease carrying droplets.

### Requirements

To inform the development process of the LeVe system, our target requirements were defined as delivery of CPAP at a mean pressure of 10 cm H_2_O (1000 Pa). The flowrate required to maintain positive pressure is not set *a priori*, but determined to ensure the pressure remains positive for all parts of the breathing cycle, nevertheless a typical guide value of 60 L/min provided an initial starting point for a suitable flow [19]. Within this CPAP regime, the system should achieve a minimum 40% FiO_2_ using an oxygen flow rate of 5L/min (under standard conditions) and at modest delivery pressures [19]. For context, widely used oxygen concentrators such as the Phillips Everflow have outlet pressures of ca. 0.35 barg [20].

The heterogenous nature of healthcare settings means that adoption of a specific technology requires local assessment against availability of parts, manufacturing facilities and healthcare services. To support this, a wider discussion of overall system requirements is included within this work.

### Technical Approach

The LeVe CPAP Blower system is based on the premise that a category of Brushless DC Current (BLDC) ‘fan blowers’ typically used for thermal management in electrical equipment inherently have the flow dynamics required for provision of CPAP, in general providing relatively high flow rates at low but stable pressures. We propose a design compatible with an oxygen concentrator which intrinsically requires an oxygen efficient solution. Based on frugal engineering principles, the design centres around an appropriately specified, single electric fan-blower without the need for control features of more complex fan-based CPAP systems. The resultant breathing circuit also minimizes the number of parts required for effective CPAP since careful choice of the fan and the use of a simple expiration port means that no PEEP valve is required.

We examined a range of BLDC fans capable of generating pressures in the range of 13–20 cm H_2_O (∼1300 – 2000 Pa) at flow rates of 100 L/min, thus ensuring the flow is greater than the peak inspiratory flow rate to maintain continuous positive pressure through the breathing cycle. Two multinational manufacturers (selected to help ensure supply-chain availability) supply fan models which fall within these criteria (*CUI Devices* CBM-979533B-168, *Sanyo Denki San Ace* B97 9BMB and B97 9BMC). Note that the fan characteristics limits the maximum system pressure to the zero-flow pressure providing a degree of safety for the patient with correct fan selection. Of these possibilities, we selected the *Sanyo Denki San Ace* B97 9BMC to accommodate higher flow rates.

Figure 3 shows how a fan-blower based system can offer a range of CPAP pressures through modulation of the fan speed. A breathing simulator, described in Section 4, was used to obtain measures across two representative breathing cycles. The characteristics show that the voltage input to the fan controls the overall fan speed and consequently the static pressure generated within the system. This offers the ability to adjust the mask pressure by using a voltage regulator to supply a variable supply voltage to the fan. A working range can be defined between the maximum operating speed and the point at which the fan cannot meet the required flow rates. In this instance, below 7V the mask pressure for the 500ml breathing cycle is negative; under such a situation the instantaneous peak inspiratory flow rate is higher than the maximum flow supplied by the fan, so a positive pressure cannot be maintained.

**Figure 3.**
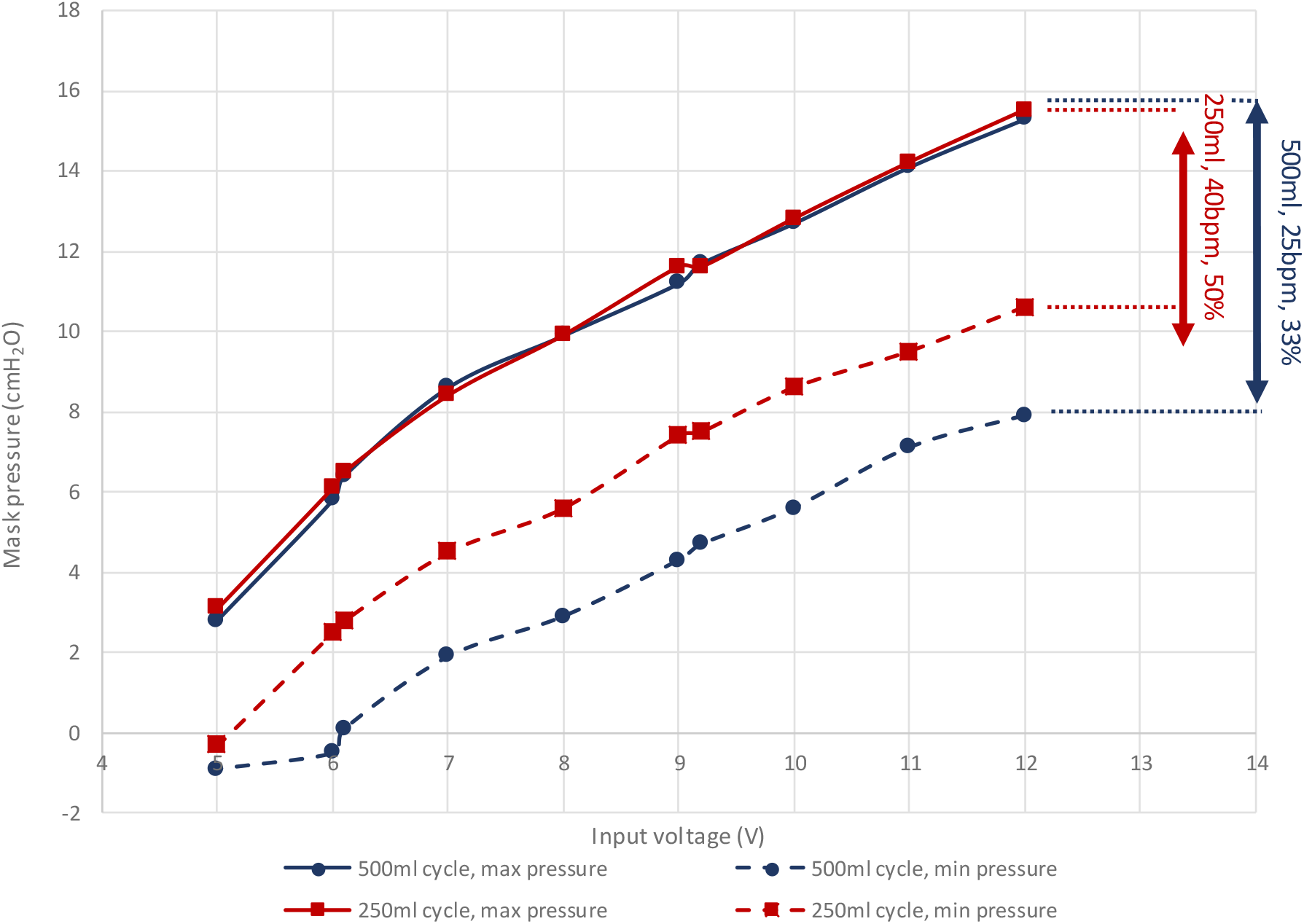
The pressure response of the LeVe Blower with varying supply voltage across different breathing cycles

### System Design

The complete LeVe CPAP system was developed after selection of the blower-fan to provide a robust package appropriate for use in low-resource contexts by trained medical practitioners. The system is designed around the fan blower, combined with a standard breathing circuit, in the configuration shown in Figure 4. In this design, rather than controlling through a supply voltage, the fan speed is controlled through a pulse-width modulated signal, generated by a low-cost PCB, which provides a low-voltage control frequency directly to the fan. A four-way dial allows selection of nominal CPAP pressures of 5, 7.5, 10 and 12.5 cm H_2_O. Power is provided to the LeVe CPAP flow generator through a medical grade 24V powerpack, which accepts an input voltage of 80V AC to 264V AC. We note that a reduced part count can be achieved by connecting the fan directly to 24V (in this case) to give a fixed CPAP pressure of 12.5cm H_2_O. A single switch on the DC circuit allows the unit to be turned on and off. An intake filter is provided to prevent particulate material entering the fan unit. The LeVe CPAP breathing circuit integrates the LeVe flow generator, to give an oxygen efficient breathing circuit, as shown Figure 4a. The circuit was implemented under the guidance of the clinical team within the Leeds NHS Teaching Hospital Trust and Bradford NHS Teaching Hospital Trust for treatment of patients with Covid-19 and adopted nationally within the UK [16], [21]. From the LeVe flow generator, there will be a constant flow of air through the expiration port. During inspiration, a fraction of air is drawn from this flow towards the patient and this oxygen is entrained into this fraction. This ensures enrichment of just the air breathed by the patient rather than requiring the entire airflow to be enriched. Exhaled air passes out of the exhalation port, with some oxygen. This loss of oxygen represents a small inefficiency within the circuit; however, the overall oxygen efficiency is still high, particularly compared to Venturi devices.

**Figure 4.**
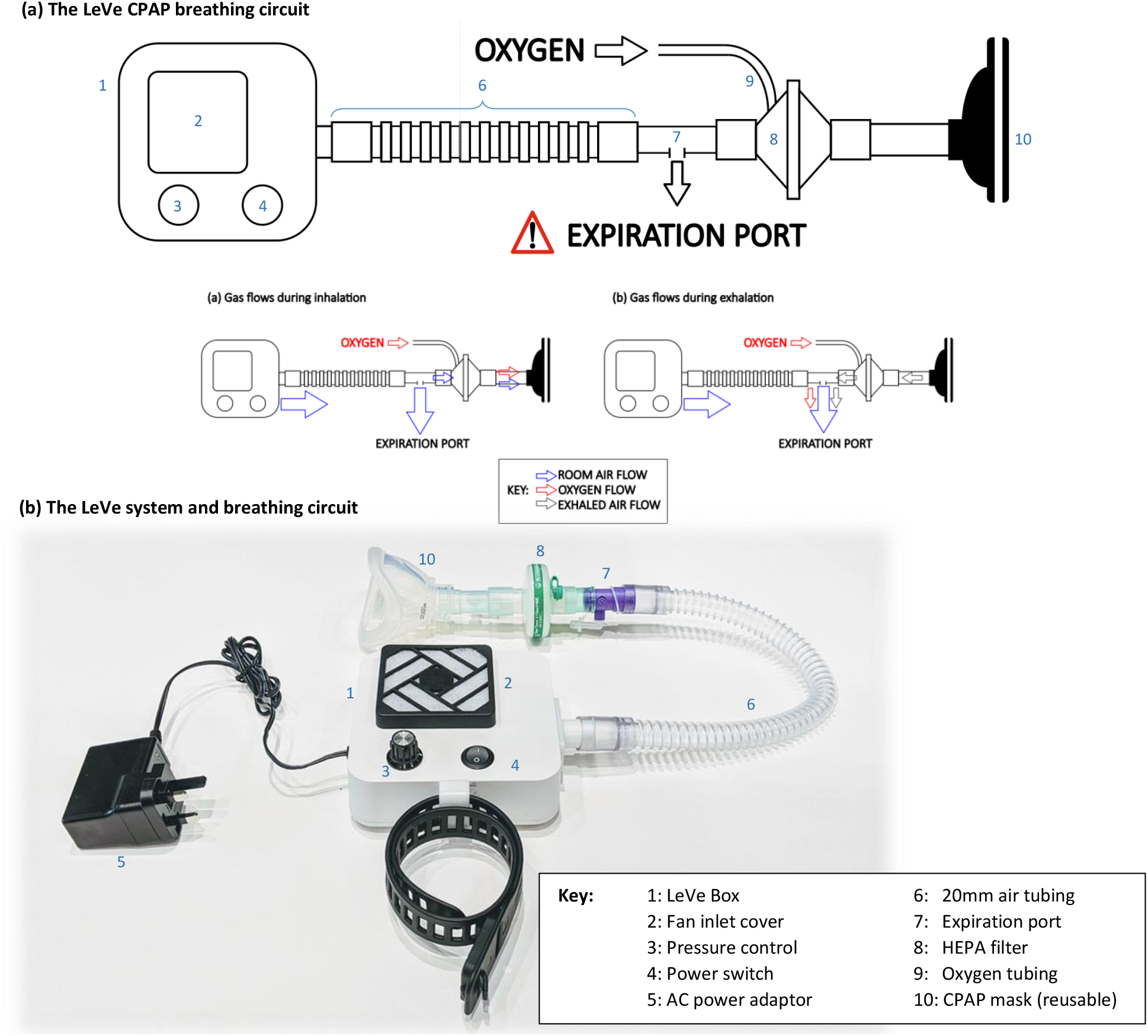
a) The LeVe CPAP breathing circuit and b) The complete LeVe system

## 3. System Evaluation

The fundamental performance characteristics of a CPAP system can be measured through: (i) the pressure at the patient mask during a breathing cycle; and (ii) the FiO2 of inspired air for a given oxygen flow rate. These aspects were investigated in controlled laboratory conditions to ensure appropriate performance prior to human use.

### Methods

To measure the flow dynamics of the system in a controlled and repeatable manner, a breathing simulator was developed. The simulator, summarised in Figure 5, consisted of a large bore pneumatic cylinder (SMC CQ2 Series, 160mm diameter) driven by a dynamic testing machine (Electro Puls E10000, Instron) which allows the cylinder piston to be moved under a pre-defined cyclic pattern, facilitating a variety of breathing cycles (see Figure 5, inset, for example) to be tested. The system enables the impact of breathing patterns to be assessed in terms of oxygen efficiency and pressure response of the circuit. 4 breathing cycles were used (Table 1) to represent a spread of respiratory cases, from slow deep breathing in healthy adults to more rapid shallow breathing associated with conditions like COVID-19 [22].

**Table 1.** Breathing cycle parameters used to evaluate the LeVe systems

| Tidal Volume (ml) | BPM | Duty Cycle (I:E) | Reference |
| --- | --- | --- | --- |
| 250 | 40 | 1:1 | Kallet |
| 500 | 25 | 2:1 | Brusasco |
| 500 | 20 | 1.5:1 | MHRA, Schneider, Wilkins |
| 500 | 25 | 1.5:1 | Baseline |

**Figure 5.**
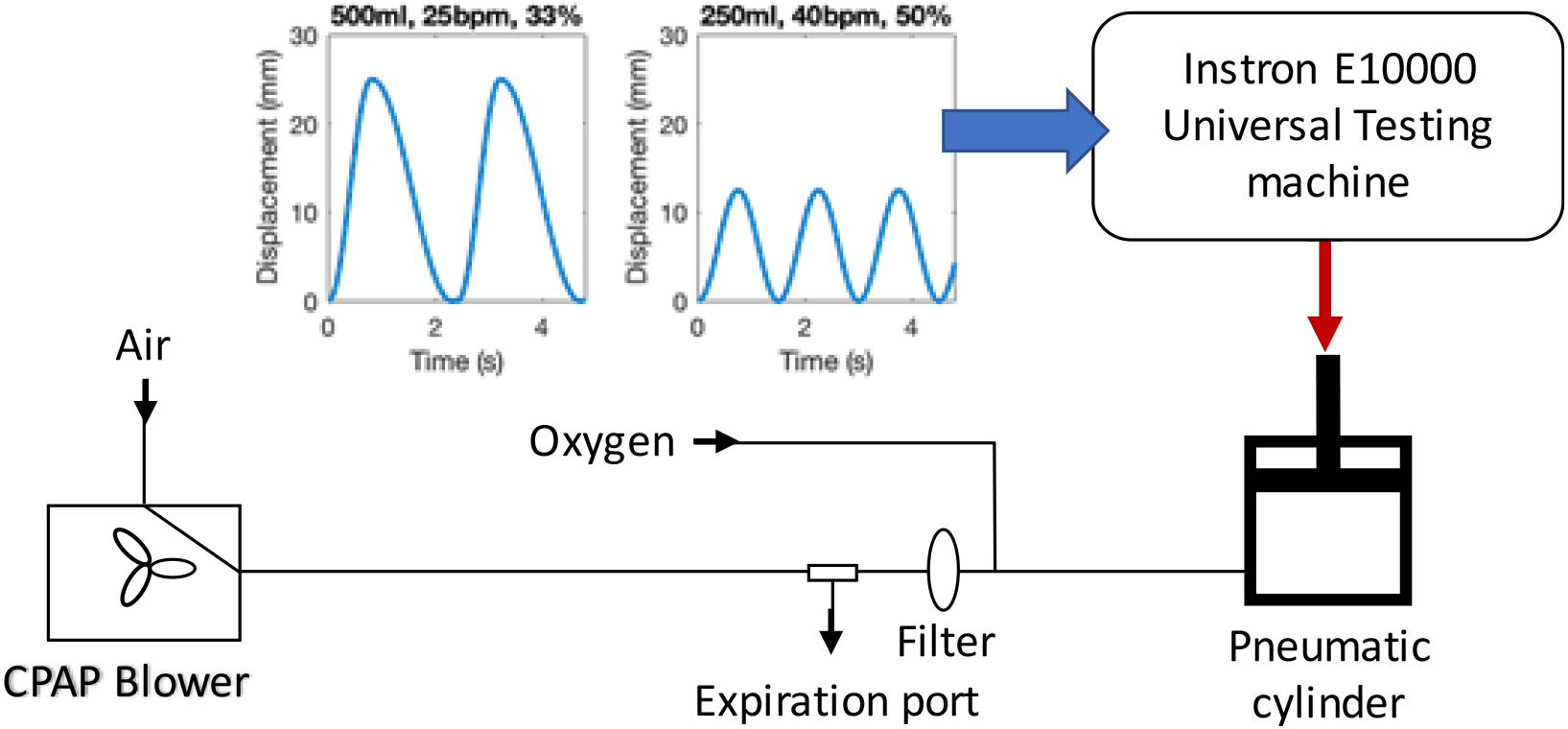
The breathing model and test configuration used to evaluate the LeVe systems

Key parameters measured within the system were mask pressure (IPSU-M12, RS), outlet flowrate of the blower (SFM3300, Sensiron), and oxygen concentration in the mask (Max-550E, Maxtec). Pressure and flowrate data was logged using a data acquisition system (cRIO, National Instruments) at 200 Hz whilst the oxygen concentration was recorded once a steady state was reached. In each configuration, measurements were averaged over 10 cycles. Oxygen was supplied using a concentrator (Drive 10 L/min DeVilbiss Healthcare). Each system was connected to the breathing simulator using standard medical-grade components.

### Results

Figure 6 shows a comparison of the pressure swing measured within the mask over 3 breaths. For all circuits, a drop in pressure is observed during inspiration with air drawn from the breathing limb, and a rise observed during expiration. This pressure swing is similar to that observed for CPAP machines, in both commercial and open-source systems [23]. The performance of the LeVe blower system is notably similar to that of the commercial CPAP system given that there is no active control. The nature of centrifugal fans such as those used here means that as the pressure within the circuit falls during inspiration, then the flow rate increases as a consequence of the fan performance characteristics.

**Figure 6.**
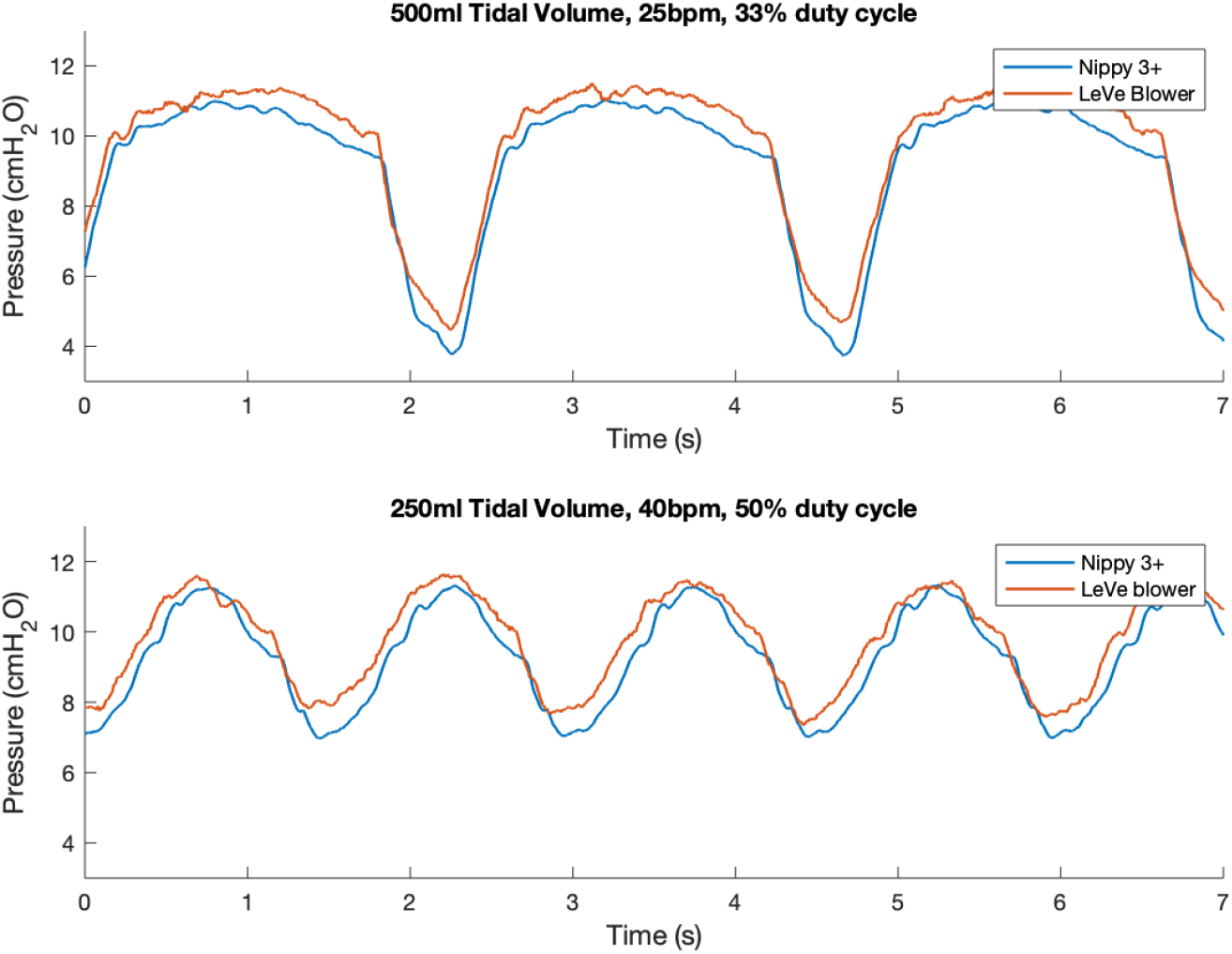
Pressure response characteristics of the LeVe systems

Figure 7 presents a summary box plot indicating the median, upper, and lower quartile as well as the maximum and minimum values. Unsurprisingly, given the observations in Figure 6, the pressure characteristics for the three systems are remarkably similar. Generally, there was a small difference as a result of the breathing pattern, with the largest range in pressure swing observed for the greatest inspiration rates, but pressures remained positive in all cases.

**Figure 7.**
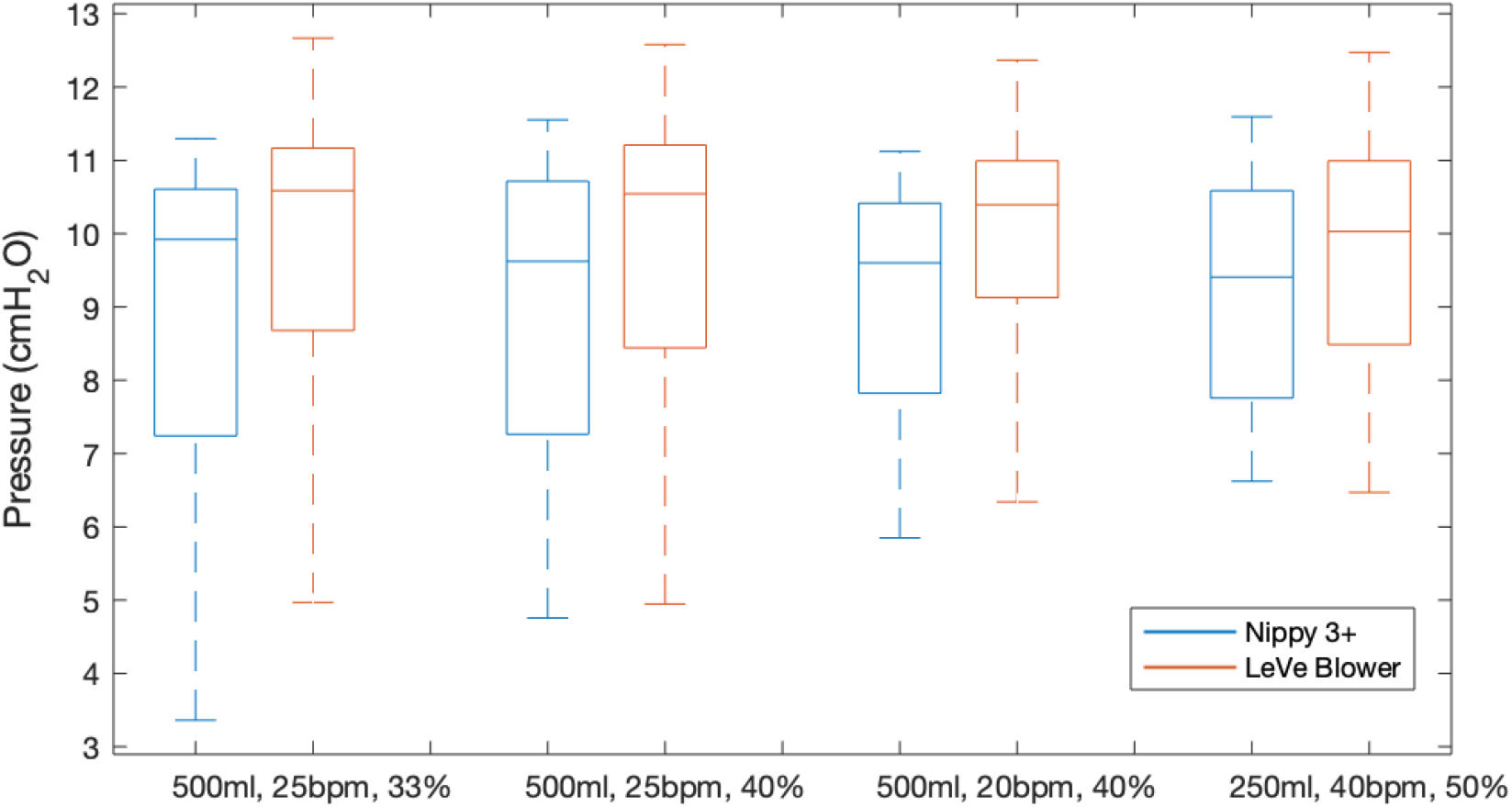
Summary of the pressure response characteristics of the LeVe system

Figure 8 illustrates the oxygen efficiency of the Leve system alongside that of the Nippy 3+. In terms of oxygen performance, the Nippy 3+ was found to have the greatest oxygen efficiency, closely followed by the Leve blower. We attribute this to the different characteristics of the fans within these systems which affects the fraction of exhaled air that is rebreathed and that which leaves the system through the expiration port. The breathing cycle has an effect on the overall performance, with lower peak inspiration rates (lower breaths per minute, smaller tidal volumes, and/or lower I:E ratios) all increasing the efficiency, due to the way the oxygen builds up in the circuit.

**Figure 8.**
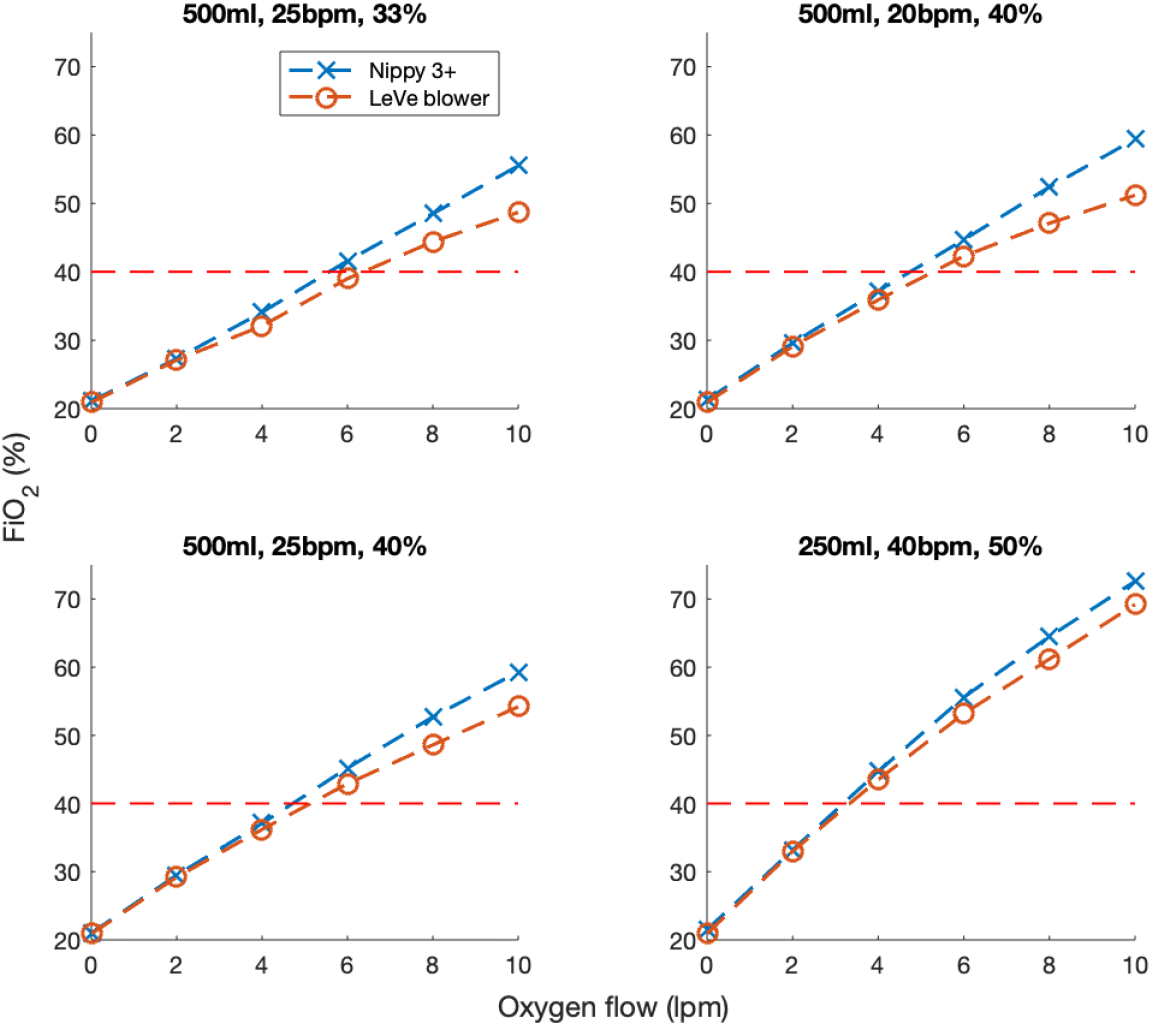
FiO2 characteristics of the LeVe breathing circuit in comparison to a sleep apnea CPAP system (Nippy 3+)

## 4. Pilot Study

A pilot study was conducted to evaluate the safety and acceptability of the LeVe CPAP Flow Generator in a group of healthy volunteers at Mengo Hospital in Kampala, Uganda. Two research questions were set:

Primary: Can the LeVe CPAP Flow Generator be used safely without inducing hypoxia or hypercapnia? Secondary: Is the LeVe CPAP Flow Generator well tolerated by users?

### Methods

This study took place in the Intensive Care Unit at Mengo Hospital, Kampala, Uganda. A sample of 10 participants was recruited, all of whom were members of staff at the hospital. All participants were provided with a written information sheet and gave informed written consent.

The inclusion criteria were:

- Staff members at Mengo Hospital
- Age 20-50 years

The exclusion criteria were:

- Current or ex-smoker
- Underlying respiratory conditions
- BMI >30
- Any contraindications from previous use of CPAP or oxygen therapy

Approval was obtained from the Mengo Hospital Research and Ethics Committee, The National Drug Authority and the Uganda National Council for Science and Technology.

The participants first trialled the mask to assess the fit. After taking a baseline reading, the LeVe CPAP Flow Generator was switched on and the pressure was stepped through the 4 settings (5 cmH_2_0, 7.5cmH_2_0, 10cmH_2_0 and 12.5 cmH_2_0). The participants’ oxygen saturation and end tidal CO_2_ level were monitored continuously throughout the study and recorded once stable at each pressure. Oxygen saturation was measured by a pulse oximeter placed on a finger, while end tidal CO_2_ concentration was estimated by a measure taken with a capnograph, with the sample line attached to a port on the face mask. Hypoxia was defined as oxygen saturations < 94% and hypercapnia was defined as ETc02 >5.7KPa. The null-hypothesis was no difference in oxygen saturation across the five groups represented by the 4 CPAP values and the baseline measurement. A one-way ANOVA was undertaken with a post-hoc assessment of pairwise evaluations using the Bonferroni Correction to account for multiple comparisons. The participants were then provided with a questionnaire which asked them to rate overall comfort, anxiety, claustrophobia, and difficulty in breathing using a Likert scale to assess each attribute.

### Results

In total, 10 participants were recruited. Mean age was 24.9 years (range 22-30 years) and 50% were female. Measured data for end-tidal CO_2_ and oxygen saturation levels is provided in Table 2. Data were recorded successfully for all participants with the exception of two readings at 5 cm H_2_O for participants 9 and 10 due to a reading error at this setting. Overall, the results demonstrate a consistent and desirable positive response in oxygen saturation levels across all participants. Similarly, end tidal CO2 falls within acceptable limits for each participant using the system.

**Table 2.**
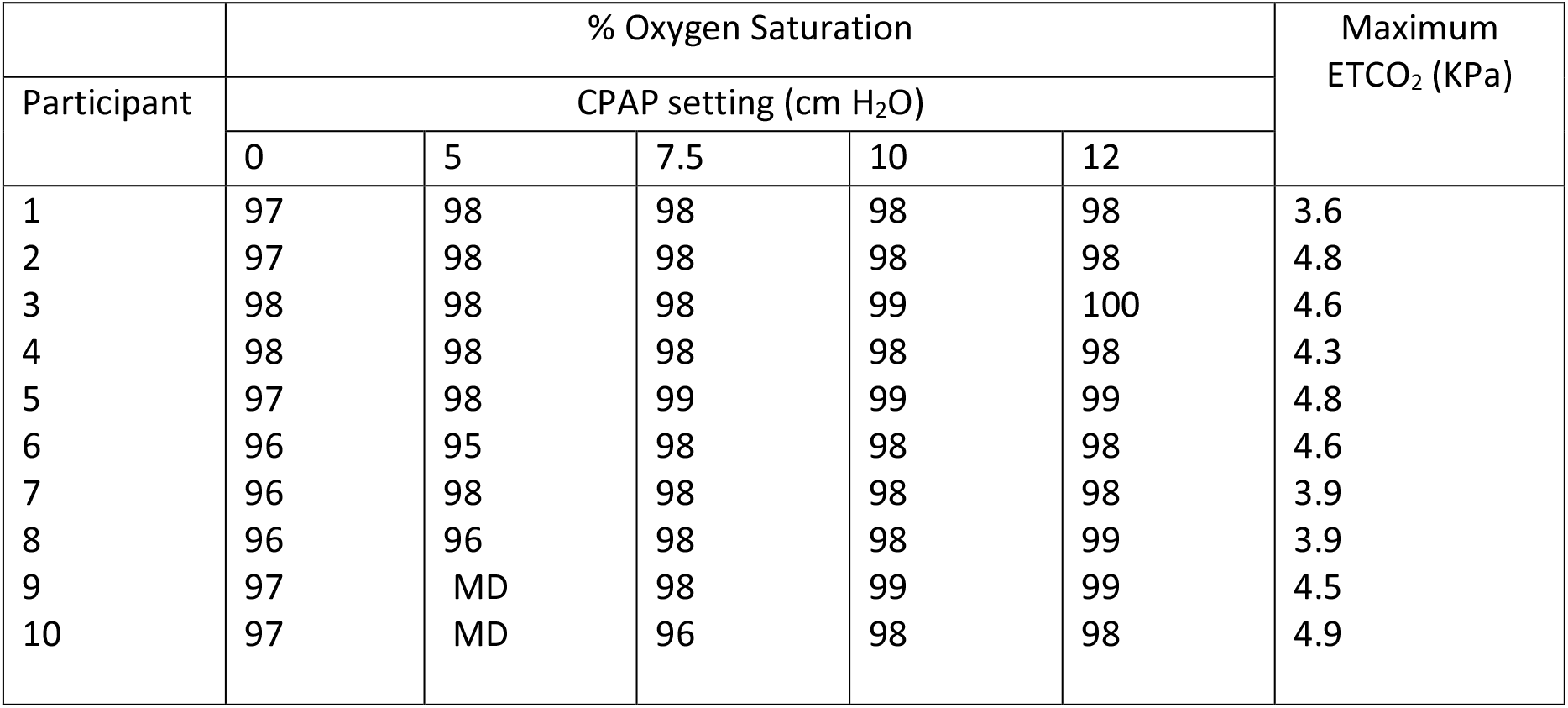
Oxygen Saturation Levels across the cohort (MD = Missing data due to reading error)

|  | % Oxygen Saturation |  |  |  |  | Maximum<br>ETCO<sub>2</sub> (kPa) |
| --- | --- | --- | --- | --- | --- | --- |
| Participant | CPAP setting (cm H<sub>2</sub>O) |  |  |  |  |  |
|  | 0 | 5 | 7.5 | 10 | 12 |  |
| 1 | 97 | 98 | 98 | 98 | 98 | 3.6 |
| 2 | 97 | 98 | 98 | 98 | 98 | 4.8 |
| 3 | 98 | 98 | 98 | 99 | 100 | 4.6 |
| 4 | 98 | 98 | 98 | 98 | 98 | 4.3 |
| 5 | 97 | 98 | 99 | 99 | 99 | 4.8 |
| 6 | 96 | 95 | 98 | 98 | 98 | 4.6 |
| 7 | 96 | 98 | 98 | 98 | 98 | 3.9 |
| 8 | 96 | 96 | 98 | 98 | 99 | 3.9 |
| 9 | 97 | MD | 98 | 99 | 99 | 4.5 |
| 10 | 97 | MD | 96 | 98 | 98 | 4.9 |

Figure 9 shows the average oxygen saturation levels for the participants as a function of the nominal CPAP setting. The ANOVA revealed a rejection of the null-hypothesis (P=0.0002) across the five groups with significant pairwise comparisons noted for both groups as denoted by CPAP values of 10 and 12 cm H_2_0 when compared with the base readings in the absence of CPAP. Thus, indicating an improvement in oxygen saturation within the participants at the two highest values of CPAP pressure. Across all participants, the maximum end tidal CO2 was below 5.7 kPa.

**Figure 9.**
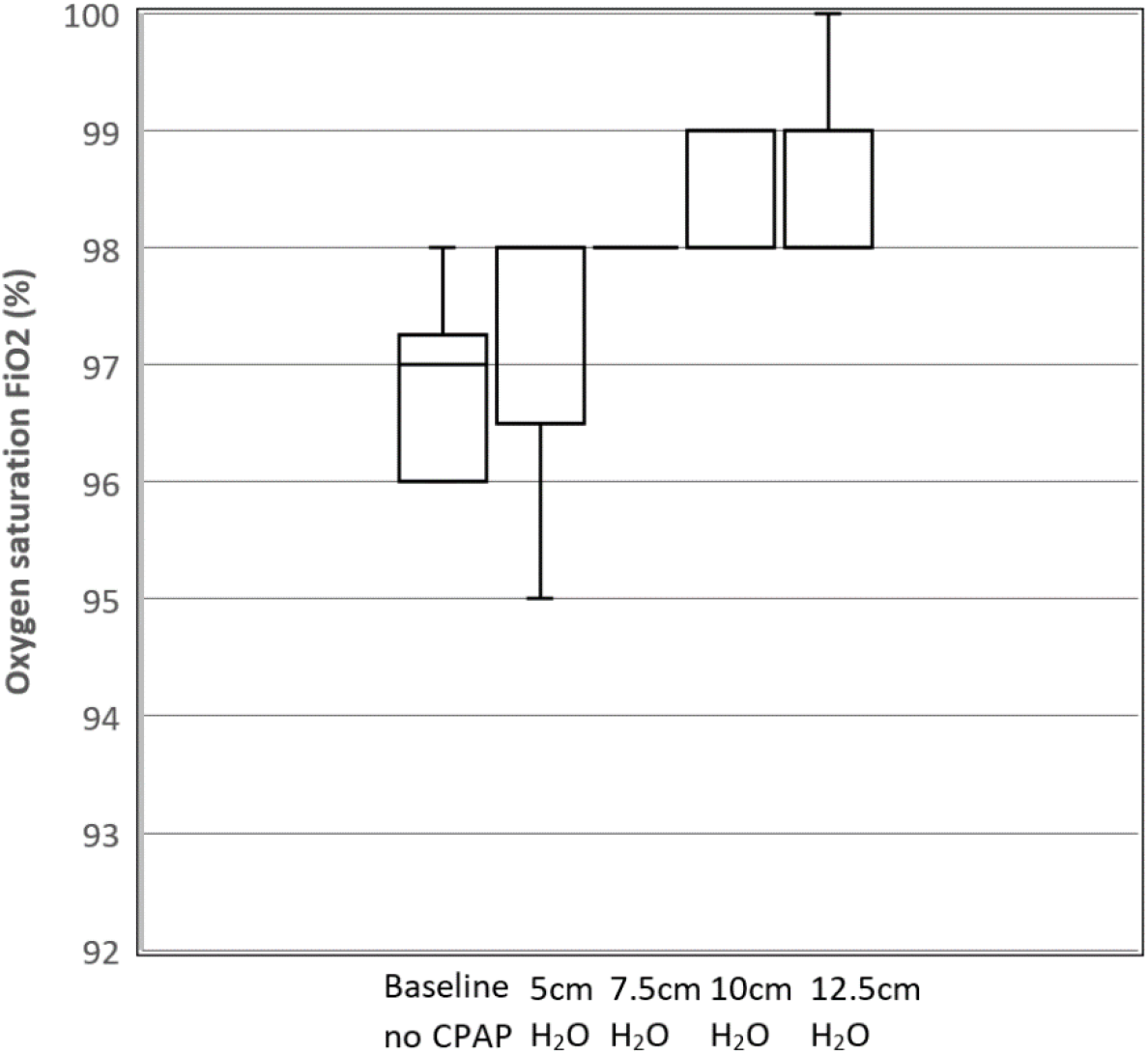
Box and whisker plot showing mean oxygen saturation levels as a function of CPAP levels for the 10 participants.

In terms of tolerability, mask comfort and overall comfort were measured on a scale of 1-5 where 1 indicated ‘not at all comfortable’ and 5 indicated ‘very comfortable’. Figure 10 demonstrates user tolerability of the device with the mean response and standard error also shown. Mean overall comfort level was 4.4. Anxiety, claustrophobia, and difficulty in breathing were then measured on a scale of 1-5 where 1 indicated ‘not at all’ and 5 indicated ‘strongly’. Mean anxiety level was 2.1, mean claustrophobia level was 1.5, and mean difficulty in breathing level was 1.9.

**Figure 10.**
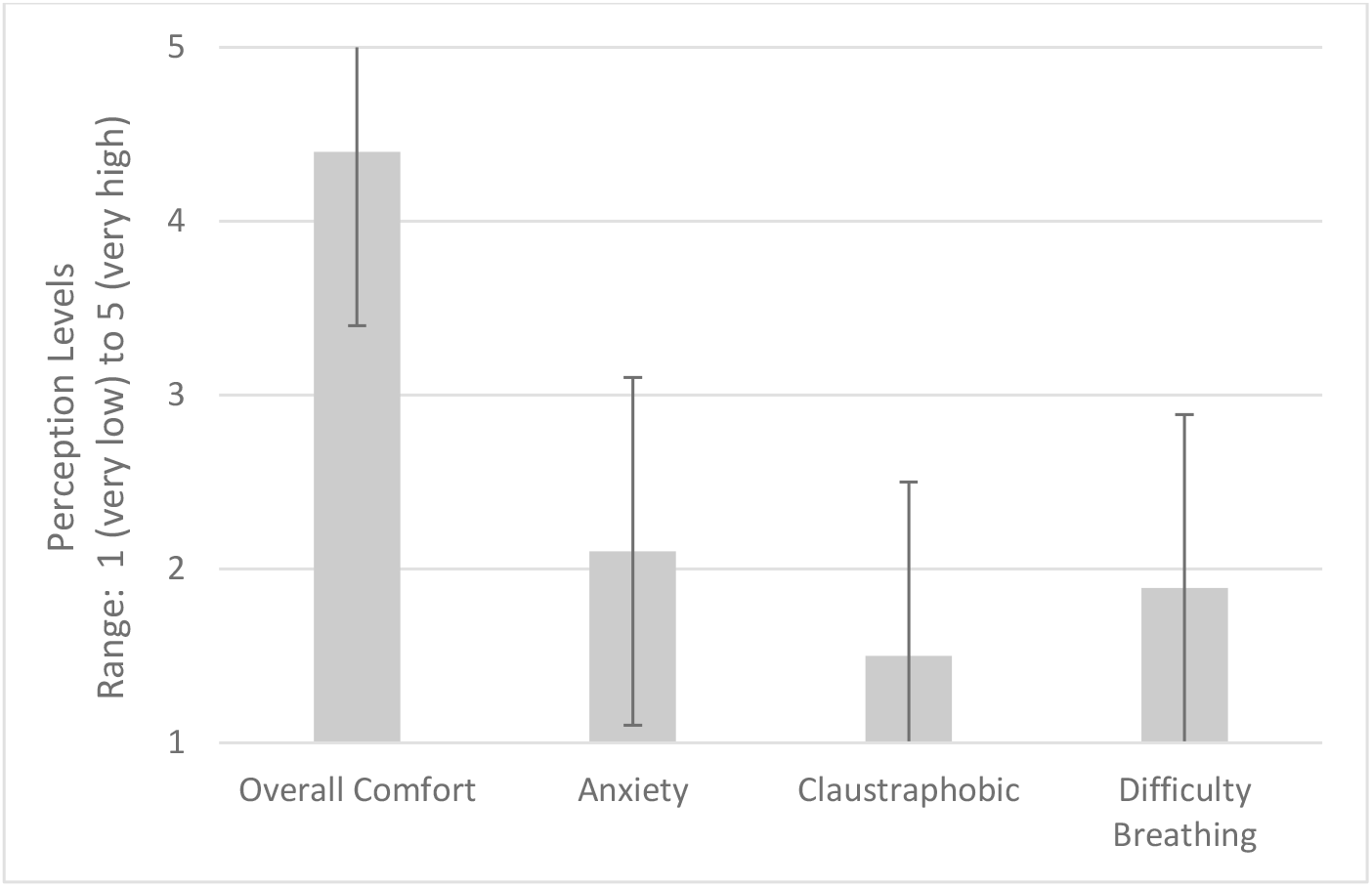
User tolerability of device. Perceptions of users during CPAP delivery using LeVe. Error bars indicate +-1 SD. 5:Very high 4:High 3:Moderate 2:Low 1:Very low

## 5. Discussion

The LeVe Blower is a simple system, deliberately developed to provide a low resource solution for the provision of CPAP ventilation in terms of oxygen requirements, power, and ease-of use whilst not sacrificing performance. Following frugal engineering principles the systems have been developed to meet a focussed set of requirements while removing extraneous functionality and thus complexity. For example, while open-sourced CPAP systems have been developed to provide low-cost alternatives to commercial systems, they fundamentally share a common approach in using microprocessor systems to regulate their output [23]. In contrast, the LeVe system uses the inherent flow characteristics of the fan to achieve comparable performance in terms of pressure and flow rate during representative breathing cycles, despite their lack of closed-loop pressure regulation. Considering resource efficiency, the LeVe system is designed to generate a pressurised airflow without the use of compressed oxygen, which is typically a limited commodity in LMICs, instead relying on more prevalent and sustainable electrical power. This approach enables the system to achieve high efficiency in the supply of oxygen-enriched air and allows oxygen delivery to be controlled independently of the desired CPAP operating pressure. In the case of LMIC context, this enables the use of oxygen concentrators as a supply.

Selection of an appropriate CPAP system for clinical use is heavily dependent on the environment, infrastructure, and resources present. Reflecting on Figure 1, our focus has been to target ‘resource-light’ solutions, to ensure that they are appropriate for LMIC contexts. This goes beyond producing a ‘low-cost’ system, instead frugal engineering emphasises the need for systems which consider manufacture, sustainable long-term use and crucially does not sacrifice performance to achieve these goals. The LeVe system is designed to operate using an oxygen concentrator to enrich the air supply. Typically, these are available in 5 or 10 L/min where the output of the latter can be split with 2 flow regulators to treat 2 patients to achieve approximately 40% FiO_2_. This provides a scalable and resource-efficient solution to cater for varying patient numbers. In contrast, operating conventional Venturi valve systems would require provision of compressed oxygen at high flow rates which is challenging to achieve in many LMICs without recourse to repeated changeover of oxygen cylinders, a practice which is both costly and demands regular maintenance support [24].

The results of the study demonstrate that in a healthy cohort, the LeVe system is safe for use and well tolerated by the participants. Measures of oxygen saturation demonstrate that the system does not induce hypoxia or reduce oxygen saturation, and in fact it may actually increase saturation levels. In a fit and well population, the significance of any increase in Sp02 with CPAP was likely to be minimal. Similarly, measures of end tidal CO_2_ show that LeVe does not cause hypercapnia. The ultimate FiO_2_ delivered by any CPAP system varies with respiratory function and is not explicitly controlled. Thus, these systems require external monitoring by a suitably qualified healthcare professional based upon the patient’s SpO2 level and vital signs in accordance with best practice (e.g., UK MHRA guidance). This places an emphasis on the need to accompany such systems with appropriate training and clinical use protocols to ensure quality of care.

Considering comfort, it is important to note that the provision of positive pressure through a face mask will inherently impact on ‘natural’ inhalation and expiration, thus may cause anxiety and discomfort, regardless of the air source. Within the scope of this study, it indicates that the air pressure and flow characteristics of the LeVe system, in particular the inherent variability which occurs during the breathing cycle, are both tolerated and appropriate for future clinical evaluation. In conjunction, the high comfort rating reported for the mask is integral to the overall experience and should be carefully selected to ensure a close but comfortable fit.

The clinical efficacy of CPAP systems has evolved rapidly during the COVID-19 pandemic [3]. Particularly in LMIC contexts, the utility of CPAP is likely to have increasing relevance to treatment of other conditions, particularly when more advanced forms of ventilation are not available. For example, CPAP provides a route to stabilise patients with acute pulmonary oedema whilst the underlying cause is being treated [25]. Similarly, based on the success of using CPAP to treat COVID-19, it could be explored for treatment of conditions like viral pneumonias or severe influenza. This need not be confined to acute settings, there is scope to explore the use of simple devices like LeVe for early presentation of COVID-19 within community settings, helping to reduce the burden on hospital admissions. Last, but not least, it is also interesting to note that while these systems were developed to target use in LMICs, there is increasing recognition of the need to innovate for value within the resource-strained healthcare systems of HICs. Often termed ‘reverse innovation’, there is also potential for the use of low-resource CPAP systems within services like the UK’s NHS [26].

## 6. Conclusions

CPAP ventilation systems provide an important treatment option for COVID-19 patients, particularly in the early stages before invasive ventilation strategies are required, to deliver oxygen-enriched air to stabilise patients until they can be escalated or de-escalated. To deliver this for the high patient numbers associated with the COVID-19 pandemic, healthcare providers require resource efficient solutions. We have shown that this can be achieved through frugal engineering of a CPAP ventilation system.

The data from the pilot study indicate that the LeVe CPAP Flow Generator is safe to use in healthy volunteers and was well tolerated by the cohort. This solution has different merits in clinical performance and efficiency to existing CPAP systems but provides resource-limited healthcare providers with a more resource-efficient solution to support flexible treatment pathways that can be rapidly deployed to reduce the burden on ICU during the COVID-19 pandemic. Beyond this immediate need, there is also evidence that CPAP can help provide much needed therapeutic benefit to address other respiratory conditions (e.g., respiratory distress syndrome) which often go unaddressed in LMICs for want of context-appropriate technology.

A complete design of the LeVe CPAP blower system is available (see supplementary materials) with our hope that this work will support the treatment of patients suffering from COVID-19 and (beyond the current pandemic) expand treatment options available to healthcare professionals targeting respiratory distress syndromes. Our ongoing work will address the clinical efficacy of using the LeVe system in patients with different respiratory conditions.

## Supporting information

Supplementary Technical Information on the LeVe System

## Data Availability

Data is available by reasonable request from the corresponding authors.

https://pactr.samrc.ac.za/TrialDisplay.aspx?TrialID=15875

## 7. Supplementary Links

Technical documentation on fan selection and system performance

## 8. Acknowledgements

We would like to thank our multidisciplinary team; Graham Brown, Sam Flint, Kevin Meloy, Mick China, James Naylor, Hardy Boocock, Tony Wiese, and the numerous healthcare professionals at Leeds and Bradford Teaching Hospitals (UK), and Mengo Hospital who have helped make this research possible. The Medical Physics teams at Leeds NHS Teaching Hospitals Trust and the Clinical Engineering Team at Cambridge University Hospitals NHS Foundation Trust for their invaluable recommendations in the development process. The Museum of Digital Art, Zurich for the provision of air sensors when none were available to buy. MedAid International for supporting the development with technical expertise and the loan of critical equipment.

## References

[1] J. Xie, Z. Tong, X. Guan, B. Du, H. Qiu, and A. S. Slutsky, “Critical care crisis and some recommendations during the COVID-19 epidemic in China,” Intensive Care Med, vol. 46, no. 5, pp. 837–840, May 2020, doi: 10.1007/s00134-020-05979-7.

[2] J. J. Marini and L. Gattinoni, “Management of COVID-19 Respiratory Distress,” JAMA, Apr. 2020, doi: 10.1001/jama.2020.6825.

[3] C. Guérin and P. Lévy, “Easier access to mechanical ventilation worldwide: an urgent need for low income countries, especially in face of the growing COVID-19 crisis,” European Respiratory Journal, vol. 55, no. 6, Jun. 2020, doi: 10.1183/13993003.01271-2020.

[4] C. Galbiati et al., “Mechanical Ventilator Milano (MVM): A Novel Mechanical Ventilator Designed for Mass Scale Production in Response to the COVID-19 Pandemic,” 2003.10405 [physics], Apr. 2020, Accessed: Jul. 13, 2020. [Online]. Available: http://arxiv.org/abs/2003.10405

[5] “Life saving breathing aid developed to keep COVID-19 patients out of intensive care.” https://www.nihr.ac.uk/news/life-saving-breathing-aid-developed-to-keep-covid-19-patients-out-of-intensive-care/24542 (accessed Jul. 13, 2020).

[6] “CAS-ViewAlert.” https://www.cas.mhra.gov.uk/ViewandAcknowledgment/ViewAlert.aspx?AlertID=103013 (accessed Jul. 13, 2020).

[7] G. W. Glover and S. J. Fletcher, “Assessing the performance of the Whisperflow® continuous positive airway pressure generator: a bench study,” Br J Anaesth, vol. 102, no. 6, pp. 875–881, Jun. 2009, doi: 10.1093/bja/aep077.

[8] D. J. Bennett, R. W. Carroll, and R. M. Kacmarek, “Evaluation of a Low-Cost Bubble CPAP System Designed for Resource-Limited Settings,” Respiratory Care, vol. 63, no. 4, pp. 395–403, Apr. 2018, doi: 10.4187/respcare.05762.

[9] M. Falk, S. Donaldsson, and T. Drevhammar, “Infant CPAP for low-income countries: An experimental comparison of standard bubble CPAP and the Pumani system,” PLOS ONE, vol. 13, no. 5, p. e0196683, May 2018, doi: 10.1371/journal.pone.0196683.

[10] R. Farré and J. M. Montserrat, “Principles of CPAP and auto-adjusting CPAP devices,” Breathe, vol. 5, no. 1, pp. 42–50, Sep. 2008.

[11] R. Tino et al., “COVID-19 and the role of 3D printing in medicine,” 3D Printing in Medicine, vol. 6, no. 1, p. 11, Apr. 2020, doi: 10.1186/s41205-020-00064-7.

[12] L. Cavallo, A. Marcianò, M. Cicciù, and G. Oteri, “3D Printing beyond Dentistry during COVID 19 Epidemic: A Technical Note for Producing Connectors to Breathing Devices,” Prosthesis, vol. 2, no. 2, Art. no. 2, Jun. 2020, doi: 10.3390/prosthesis2020005.

[13] T. Duke, D. Peel, S. Graham, S. Howie, P. Enarson, and R. Jacobson, “Oxygen concentrators: A practical guide for clinicians and technicians in developing countries,” Annals of tropical paediatrics, vol. 30, pp. 87–101, Jun. 2010, doi: 10.1179/146532810X12637745452356.

[14] R. Neighbour, R. Eltringham, C. Reynolds, and J. Meek, “Affordable CPAP in low income countries,” p. 3.

[15] H. Graham et al., “Providing oxygen to children in hospitals: a realist review,” Bulletin of the World Health Organization, vol. 95, no. 4, pp. 288–302, Apr. 2017, doi: 10.2471/BLT.16.186676.

[16] T. Lawton et al., “Reduced critical care demand with early CPAP and proning in COVID-19 at Bradford: a single centre cohort,” medRxiv, p. 2020.06.05.20123307, Mar. 2021, doi: 10.1101/2020.06.05.20123307.

[17] S. Alviset et al., “Continuous Positive Airway Pressure (CPAP) face-mask ventilation is an easy and cheap option to manage a massive influx of patients presenting acute respiratory failure during the SARS-CoV-2 outbreak: A retrospective cohort study,” PLOS ONE, vol. 15, no. 10, p. e0240645, Oct. 2020, doi: 10.1371/journal.pone.0240645.

[18] T. Weyrauch and C. Herstatt, “What is frugal innovation? Three defining criteria,” Journal of Frugal Innovation, vol. 2, no. 1, p. 1, Dec. 2016, doi: 10.1186/s40669-016-0005-y.

[19] C. Brusasco, F. Corradi, A. De Ferrari, L. Ball, R. M. Kacmarek, and P. Pelosi, “CPAP Devices for Emergency Prehospital Use: A Bench Study,” Respiratory Care, vol. 60, no. 12, pp. 1777–1785, Dec. 2015, doi: 10.4187/respcare.04134.

[20] D. Peel, R. Neighbour, and R. J. Eltringham, “Evaluation of oxygen concentrators for use in countries with limited resources,” Anaesthesia, vol. 68, no. 7, pp. 706–712, 2013, doi: 10.1111/anae.12260.

[21] P. Culmer et al., “Delivery of CPAP respiratory support for COVID-19 using repurposed technologies,” Respiratory Medicine, preprint, Apr. 2020. doi: 10.1101/2020.04.06.20055665.

[22] L. Gattinoni et al., “COVID-19 pneumonia: different respiratory treatments for different phenotypes?,” Intensive Care Med, vol. 46, no. 6, pp. 1099–1102, Jun. 2020, doi: 10.1007/s00134-020-06033-2.

[23] O. Garmendia et al., “Low-cost, easy-to-build noninvasive pressure support ventilator for under-resourced regions: open source hardware description, performance and feasibility testing,” European Respiratory Journal, vol. 55, no. 6, p. 2000846, Jun. 2020, doi: 10.1183/13993003.00846-2020.

[24] R. Inglis, E. Ayebale, and M. J. Schultz, “Optimizing respiratory management in resource-limited settings,” Curr Opin Crit Care, vol. 25, no. 1, pp. 45–53, Feb. 2019, doi: 10.1097/MCC.0000000000000568.

[25] A. Gray, S. Goodacre, D. E. Newby, M. Masson, F. Sampson, and J. Nicholl, “Noninvasive Ventilation in Acute Cardiogenic Pulmonary Edema,” New England Journal of Medicine, vol. 359, no. 2, pp. 142–151, Jul. 2008, doi: 10.1056/NEJMoa0707992.

[26] M. Skopec, H. Issa, and M. Harris, “Delivering cost effective healthcare through reverse innovation,” BMJ, vol. 367, p. 6205, Nov. 2019, doi: 10.1136/bmj.l6205.

