## Supplementary Technical Information on the LeVe System for "The LeVe CPAP System for oxygen-efficient CPAP respiratory support: Development and pilot evaluation"

Design Development

#### Introduction

Constant positive airway pressure (CPAP) has many clinical application, particularly in light of the current COVID-19 pandemic, where it is being used as an early treatment option, reducing the need for mechanical ventilation. As well as venturi devices, which are driven by compressed air or oxygen, CPAP can be provided by dedicated machines such as the Nippy 3+ sleep apnea machine. Figure 1 shows the typical breathing circuit which should be used in conjunction with a CPAP blower; the injection of oxygen near the mask can be used in conjunction with CPAP for even greater therapeutic benefit. Addition of oxygen closer to the patient is generally more efficient in the use of oxygen. In this breathing circuit, the inclusion of the expiration port is important since it provides an additional outlet for exhaled gases and reduces rebreathing.

Although sleep apnea machines are commercially available, they are often prohibitively expensive for use in low to middle-income countries (LMICs) and demand during the COVID-19 pandemic is far outstripping supply. This document will outline a frugal engineering approach to creating a CPAP blower device using readily available parts.

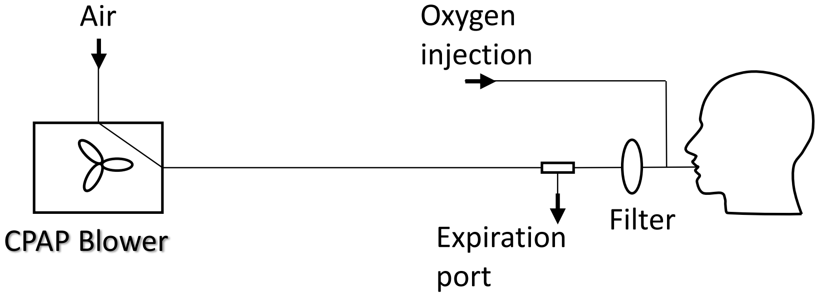


*Figure 1: Typical breathing circuit for a CPAP blower*

#### Testing rig

Testing was performed on a mechanical lung simulator which allowed for the dynamics of the system to be measured in a controlled, repeatable way. The simulator, shown in Figure 2, consisted of a large bore pneumatic cylinder (SMC CQ2 Series, 160mm diameter) driven by a dynamic testing machine (Electro Puls E10000, Instron) which allows the cylinder piston to be moved under a pre-defined cyclic pattern, allowing a variety of breathing cycles (see Figure 2, inset, for example) to be tested. Such a system allows for the impact of breathing patterns to be assessed in terms of oxygen efficiency and pressure response of the circuit. Key parameters measured within the system include mask pressure (IPSU-M12, RS), outlet flowrate of the blower (SFM3300, Sensiron) and oxygen concentration in the mask (Max-550E, Maxtec). Pressure and flowrate data were logged using a data acquisition system (myRIO, National Instruments) at 200 Hz whilst the oxygen concentration was recorded once steady-state was reached. Both bottled oxygen (99.5% O2 BOC) and oxygen via a concentrator (Drive 10L/min DeVilbiss Healthcare) was available with the latter used throughout except for the performance measurements of the oxygen driven venturi circuit.

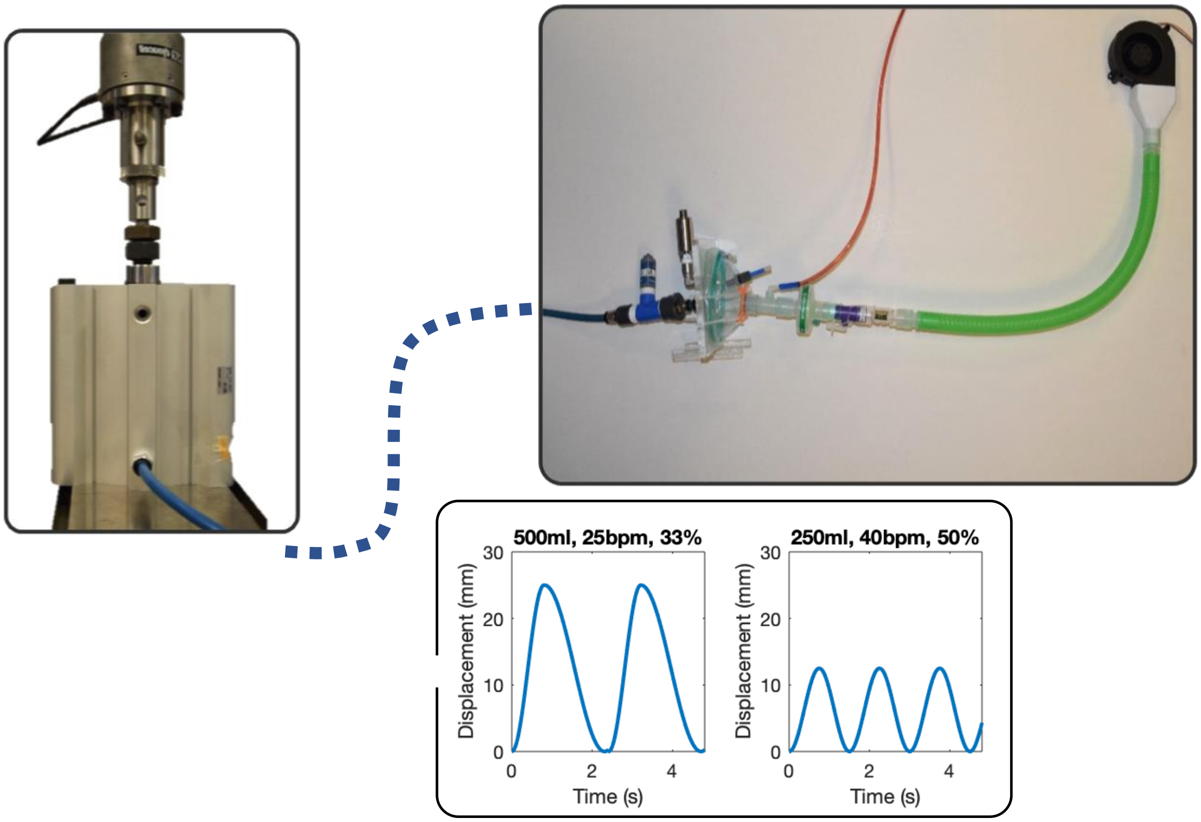


*Figure 2: Mechanical lung simulator connected to the breathing circuit. Inset: typical breathing cycles used for testing.*

### Design and part selection

#### Blower

Small fans are often used to cool electrical components: these are compact, have long lifespans and output high flow rates which make them ideal candidates for supplying the airflow in the LeVe blower. Initial testing revealed that it is possible to use these fans in an open loop system, without the need for feedback. The performance of these fans is characterized by pressure-airflow curves: the higher the downstream pressure, the lower the flow rate and vice-versa. Therefore, as the patient inhales, the downstream pressure decreases and the outputted flow rate from the fan automatically increases. Likewise, when the patient exhales, the downstream pressure increases and the flow rate in the fan reduces. It is these properties which allow the fans to maintain positive end-expiratory pressure (PEEP) throughout the duration of a patient's breathing cycle.

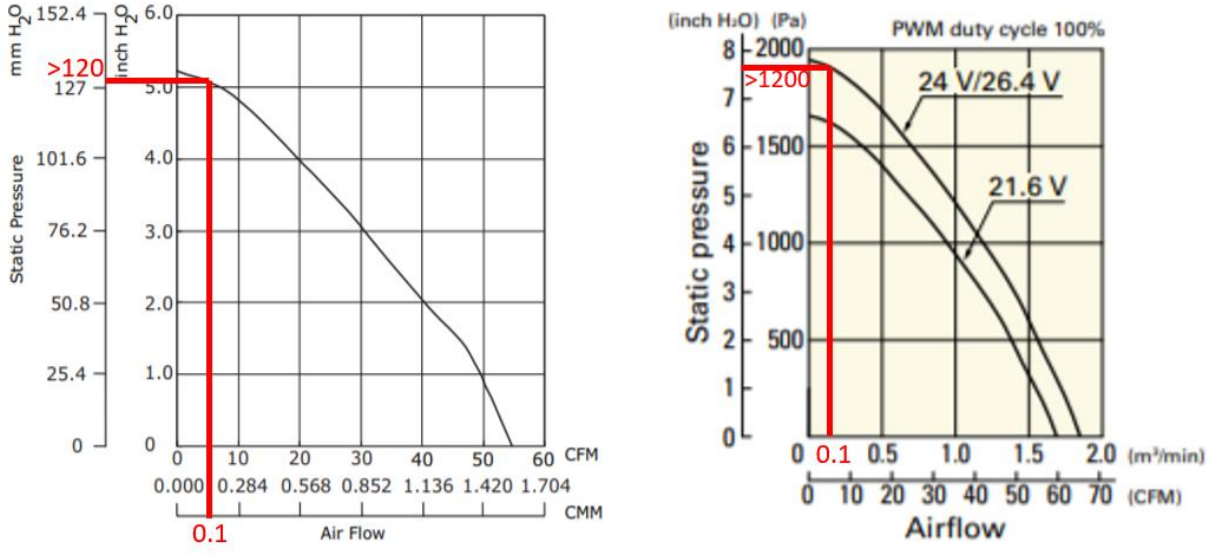


*Figure 3: Typical airflow-performance curves for a computer fan. Left shows performance for the CUI Devices model CBM-979533B-168, right shows performance for the Sanyo Denki B97. Red lines show performance criteria for fan selection.*

Typical fan performance curves are shown in Figure 3 and can be used to properly select which fan should be used. When outputting a flow of 100lpm the fan should be able to maintain a static pressure around 2cm$H_2O$ higher than required to account for any pressure losses in the medical tubing or possible voltage drops in the control system. In this case the desired maximum pressure is 10cm$H_2O$, so a maximum pressure of 12cm$H_2O$ (~1200Pa) is required. Both fans shown in the figure above would be suitable for this application – at 100lpm (0.1$m^3min^{-1}$) they produce over the required 12cm$H_2O$ (1200Pa). The ability to output a flow rate of 100lpm at this pressure is important since this is the typical peak inspiratory flow rate for an adult, therefore the fan should be able to support this flow during inspiration to maintain a positive pressure.

Another important feature to note is the driving voltage: typically, fans are either driven by a 12V or 24V DC power supply. Higher voltage fans will typically draw less current and therefore have a lower risk of overheating the electrical components, therefore these are preferable over the 12V variants. Lastly, the desired control method should be considered. Fans such as the CBM-979533B-168 (CUI Devices) can only be controlled by varying the input voltage. A variety of DC voltage controllers are available for this application. Alternatively, fans such as the B97 (Sanyo Denki) can be controlled with pulse width modulation (PWM). This is generally the preferred method of control since there are fewer losses.

#### Fan adapter

Most computer fans have a rectangular output, so a converter is necessary to attach the fan to the standard 22mm diameter medical tubing. This can either be 3D printed or machined. Due to the poor tolerances of the outer dimensions of the blower (±1mm), the fit may be quite loose – it is therefore recommended to seal the adapter to the blower either using medical grade silicone sealant or heat shrink. Adhesives are not recommended for this application since they may cause harmful fumes to be blown to the patient.

#### Enclosure

There are many available enclosures, which are normally used for electronics. They are generally made from molded ABS and available in a variety of sizes – the minimum dimensions for this application are around 150mm x 150mm x 40mm, depending on the complexity of the final product. For this device, a Takachi electric enclosure[1] was selected as a suitable candidate.

#### Control

A reliable control system was required to ensure that the fan could accurately maintain the desired mask pressure. Several possible options were considered to control the speed of the fan:

- PWM control (Via 555 timer)
- Variable voltage control (Via 24v PWM signal)
- Fixed voltage control (Via voltage regulator)

PWM control via a 555 timer circuit was selected (See supplemental material for details) as this allowed variable control of the mask pressure allowing the same system to be used for both adults and children. It also allowed the full range of the fan to be used as the variable voltage control system had a 0.8v loss across it which limited the maximum mask pressure that could be achieved.

#### Other considerations

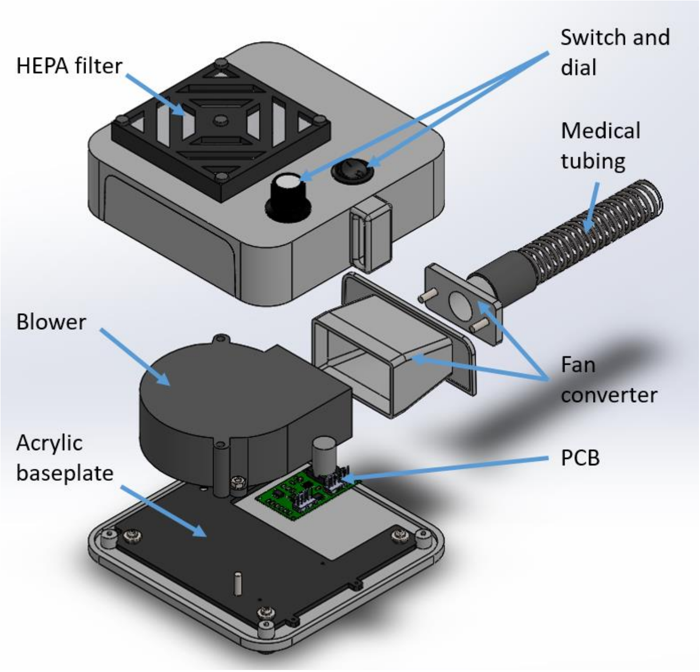


*Figure 4: Exploded view of the blower assembly with the main components annotated*

The blower and PCB are both bolted onto an acrylic baseplate which is then secured to the bottom of the enclosure so that no screws are visible from the outside of the box. The fan converter is made from two pieces which are bolted to the side panel of the box – this is then held onto and sealed around the fan with electrical heat shrink. There is a medical-grade HEPA filter on the top of the enclosure to filter the air

[1] https://uk.rs-online.com/web/p/products/1748939/

as it is drawn into the fan. Lastly, there is an on/off switch and a dial which allows for the level of CPAP to be selected.

#### Performance

The performance of the blower closely matches that of the Nippy 3+ sleep apnea machine, as shown in Figure 6: Comparison of mask pressure against time for the LeVe blower, LeVe venturi and Nippy 3+ when set at 10cmH20. In order to set the voltage (or duty cycle if using PWM control) required for a specific positive airway pressure (PAP), the blower was connected to the testing rig without a breathing cycle running. Then, the voltage (or duty cycle) required to maintain the desired PAP is recorded – this will be the value needed when a breathing cycle is added. This is shown graphically in Figure 5, where the 'Static data' shows pressure values with no cycle. It is interesting to note in this figure that for both cycles the maximum mask pressure is the same, whereas the minimum pressure is lower on the cycle with a greater peak inspiratory flow rate.

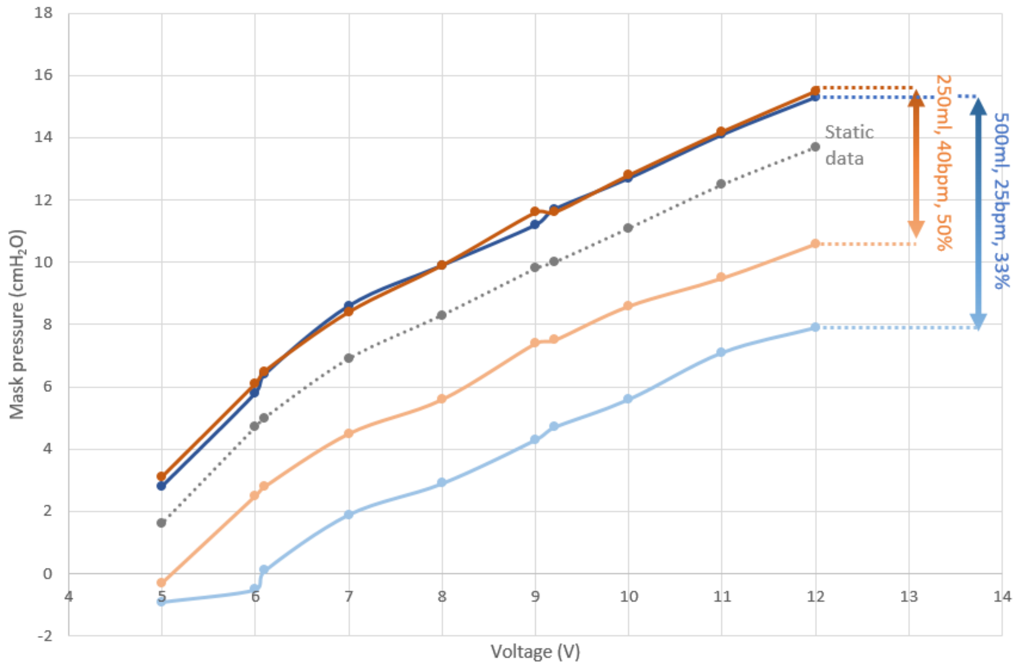


*Figure 5: Mask pressure against voltage for the CUI devices fan.*

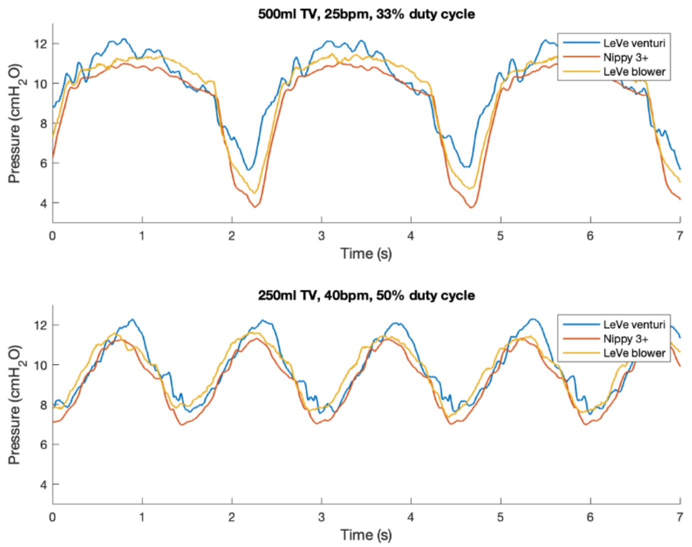


*Figure 6: Comparison of mask pressure against time for the LeVe blower, LeVe venturi and Nippy 3+ when set at 10cmH20*

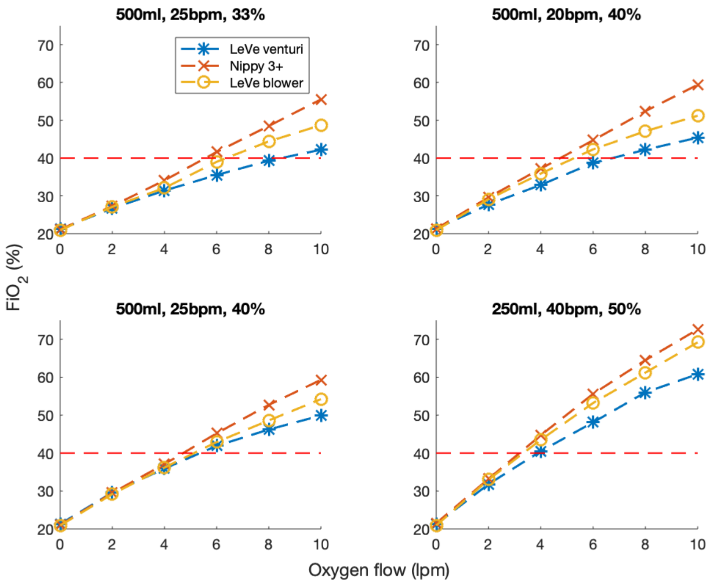


*Figure 7: $FiO_2$ performance for the LeVe venturi , LeVe blower and Nippy 3+ when set at 10cmH20*

$FiO_2$ performance, shown in Figure 7: $FiO_2$ performance for the LeVe venturi , LeVe blower and Nippy 3+ when set at 10cmH20, is similar to that of the Nippy 3+. As with Nippy and LeVe venturi, it varies with breathing cycle as the oxygen flows in near the mask rather than being mixed upstream as it is in conventional venturi devices. It is interesting to note that the LeVe blower performs better than the LeVe venturi system – this is due to the rebreathing in the circuit. Generally, the blower system has a greater amount of rebreathing, therefore the oxygen-rich air which the patient exhales is re-used, making it overall a more oxygen efficient system. In comparison, in the venturi circuit, the majority of the oxygen-rich exhaled air is passed out through the PEEP. This is shown schematically in Figure 8. The slightly improved performance of the Nippy 3+ compared to the LeVe blower is not fully known, but could be due to the Nippy having a greater modulation of flow throughout the breathing cycle compared to the blower, and thus a greater degree of rebreathing is occurring.

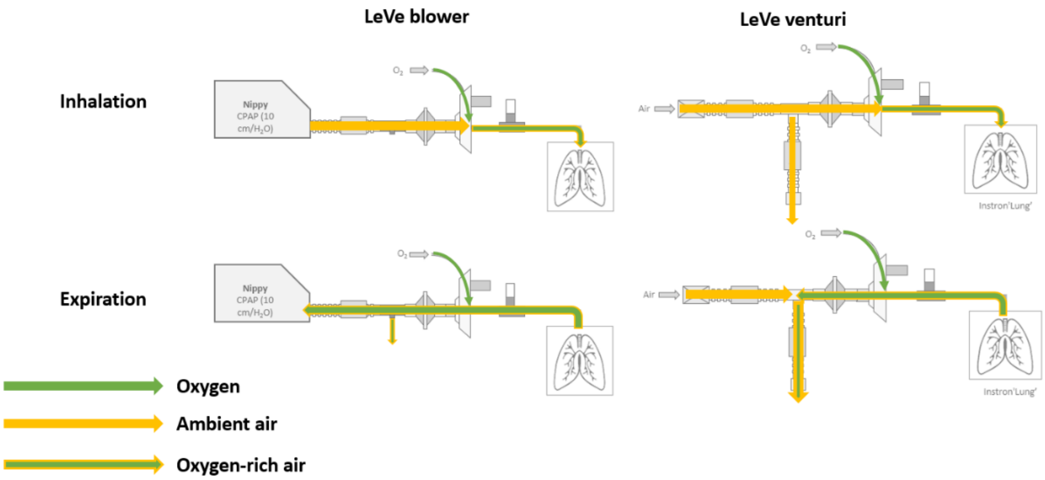


*Figure 8: Comparison of the air and oxygen dynamics in the LeVe blower and LeVe venturi systems.*
